# A next generation CRISPR diagnostic tool to survey drug resistance in Human African Trypanosomiasis

**DOI:** 10.1101/2024.09.15.24313552

**Authors:** Elena Pérez Antón, Annick Dujeancourt-Henry, Brice Rotureau, Lucy Glover

## Abstract

The WHO aims to eliminate the *gambiense* form of human African trypanosomiasis (HAT) by 2030. With the decline of reported cases, maintaining efficient epidemiological surveillance is essential, including the emergence of drug-resistant strains. We have developed new highly specific diagnostic tools using Specific High-Sensitivity Reporter Enzymatic UnLOCKing (SHERLOCK) technology for monitoring the presence of drug-resistant genotypes that (1) are already circulating, such as the AQP2/3_(814)_ chimera providing resistance to pentamidine and melarsoprol, or (2) could emerge, such as *Tb*CPSF3 (N^232^H), associated to acoziborole resistance in lab conditions. The melarsoprol - pentamidine*AQP2/3_(814)_* SHERLOCK assay detected RNA from both cultured parasites and field isolated strains from gHAT patients in relapse following treatment. The acoziborole *CPSF3_(SNV)_*SHERLOCK assay discriminated between wild-type *CPSF3* RNA and *CPSF3* with a single A-C mutation that confers resistance to acoziborole *in vitro*.

## Introduction

Human African trypanosomiasis (HAT), or sleeping sickness, is one of the 21 conditions identified as a neglected tropical diseases by the World Health Organization (WHO) ^1^. Caused by an infection with the extracellular protozoan parasite *Trypanosoma (T) brucei (b) gambiense* (gHAT) in West and Central Africa or *T. b. rhodesiense* (rHAT) in East and Southern Africa. HAT infections follow a typical clinical pattern, initiating with intermittent fever and lymphadenopathy during a first step of blood and lymph infection (stage 1) and advancing to severe neurological symptoms when the parasites invade the cerebrospinal fluid (stage 2), and ultimately death if untreated. The major difference in disease progression between *T. b. gambiense* and *T. b. rhodesiense* is that the former is chronic lasting over several months to years and the latter acute, lasting for several weeks. There is currently no prophylactic drug available for HAT, the approved chemotherapies depend on the parasite species and the stage of the disease^1^. For gHAT, children under the age of 6 or less than 20 kg are, as a first choice, treated with pentamidine for stage 1 or NECT for stage 2. Patients older than 6 years old and more than 20 kgs are treated with fexinidazole at stage 1, or with fexinidazole (if white blood cell count in the cerebrospinal fluid is lower than 100/μL) or NECT at stage 2 ^2^. If relapse is detected, patients are treated with NECT. For rHAT, children under the age of 6 or less than 20 kg are treated with suramin as a first choice for stage 1 or melarsoprol for stage 2, and if relapse is seen, fexinidazole may be given for compassionate use ^2^. Patients older than 6 years old and more than 20 kgs are treated with fexinidazole for both stage 1 and 2, yet if relapse is detected, patients are treated with suramin or melarsoprol ^2^. Both suramin and pentamidine require prolonged intravenous administration, melarsoprol causes encephalopathic syndrome that is fatal in up to one in ten patients and NECT involves a long and complex dose regime with intravenous eflornithine alongside oral nifurtimox over the course of two weeks at hospital ^3^. These drugs present either with side effects or, are logistically challenging to administer. Accurate diagnosis and staging of the disease are key for the selection of the appropriate drug treatment, but diagnosis is based on a tedious algorithm, including lumbar puncture as a confirmatory test for the most advanced stage of gHAT^4^. The high toxicity of some of these drugs, especially melarsoprol, as well as the complex administration of NECT make access to treatment challenging and have prevented implementation of mass treatment as a method of combating the disease ^5^.

Fexinidazole was approved in 2019 by the European Medicines Agency ^6^, as a 10-day oral treatment regime that must be taken with food for optimal absorption, yet with treatment emergent adverse events such as vomiting and nausea increases the risk of non-compliance ^5,7^. Due to the high-risk of non-compliance leading to the ingestion of suboptimal curative doses, the possibility of relapse is of concern – especially as it may occur late, up to 24 months post- treatment ^5^, which could increase the risk of the emergence of drug-resistance. Resistance to fexinidaxole is readily generated under laboratory conditions ^8^ and occurs through a similar mechanism as that to nifurtimox, via mutation of the nitroreductase (NTR) gene ^8^. Therefore, there already exists the potential that previous use of nifurtimox in the field may have already led to the emergence of fexinidazole resistant parasites ^9^. Fexinadazole is currently the only oral drug approved for treatment of both stage 1 and 2 gHAT and rHAT ^2,5^. Drug resistance is not uncommon in the treatment of HAT, melarsoprol resistance emerged as early as 1970 and was widespread by 1990 ^10,11^, while resistance to both eflornithine and nifurtimox can be generated *in vitro* ^3^.

In a context where the therapeutic arsenal for treating HAT is limited, the emergence of drug resistance is possible where a selective pressure to survive is applied to parasites, placing both existing and potential future drug treatments at risk. Pentamidine and melarsoprol share the same mechanism of entry into the parasite cell through the adenosine transporters and the aquaglyceroporin channels (AQPs) ^10,12^. Among the three aquaglyceroporin genes of *T. brucei* (*TbAQP1-3*) ^13^, the deletion of the *AQP2* locus is related to the melarsoprol-pentamidine cross-resistance in bloodstream-forms, increasing their effective concentrations (EC_50_) by 2- to 15-fold, respectively ^12^. Several distinct mutations in the *AQP2-AQP3* genes have been associated with relapse of HAT after treatment with melarsoprol in patients from the Mbuji-Mayi region of the Democratic Republic of the Congo ^14,15^ and the Mundri county of South Sudan^14,16^ . One specific chimera identified, a chimera containing the first 813 bp from *AQP2* and the last 126 bp from *AQP3*, here termed AQP2/AQP3_(814)_, ^15^, was associated with cross-reactivity to melarsoprol and pentamidine ^14^.

Although not a frontline drug yet, acoziborole promises to be key in the efforts to eliminate HAT. Currently in phase III clinical trials, acoziborole could be approved for the treatment of both stages of gHAT by 2026 ^17^, it is a single oral dose benzoxaborole derivative that shows high efficacy and safety. Its approval could eliminate the need for routine lumbar puncture and would allow the treatment of parasitology-negative suspects, as well as making treatment more accessible to patients living in remote areas ^17,18^. The drug molecule binds to the active site of the Cleavage and Polyadenylation Specificity Factor 3 (CPSF3), which is involved in trypanosome mRNA processing ^19^, inhibits polypeptide translation and reduces endocytosis of haptoglobin-hemoglobin ^20^. However, resistance to acoziborole can be generated *in vitro*, through editing a single nucleotide in the *CPSF3* sequence ^21^.

The CRISPR based diagnostic - Specific High-sensitivity Enzymatic Reporter unLOCKing (SHERLOCK) - first amplifies nucleic acid using recombinase polymerase amplification (RPA), which is then combined with the Cas13a nuclease, for target recognition via specific guides, and a fluorescent reporter linked to a quencher by nucleotides. Target sequence recognition activates Cas13a’s ‘collateral effect’ of promiscuous ribonuclease activity. So far, SHERLOCK diagnostic assays have been developed for two protozoan parasites, African trypanosomes^22^ and *Plasmodium*^23^. SHERLOCK is capable of detecting a few attomoles (10^-18^ moles) of nucleic acids in a sample and its specificity was demonstrated by distinguishing between closely related Zika and dengue viruses in clinical isolates, or mutant strains with unique SNPs ^24–26^. Here, we developed highly specific SHERLOCK assays that allow detection of specific drug-resistance associated mutations and that could be essential for epidemiological surveillance in the context of gHAT elimination and rHAT control.

## Methods

### Trypanosome and RNA material

*T. b. brucei* Lister 427 bloodstream form cells were cultured in HMI-11 medium with 10 % foetal bovine serum (Sigma-Aldrich) at 37 °C with 5% CO_2_. RNA extraction was carried out from culture harvest at 1x10^6^ cell/mL. RNA extraction of *T. b. brucei* cells, *T. b. gambiense* cell pellets, and human embryonic kidney (HEK) 293T cells were carried using the RNeasy Mini kit (Qiagen). Lyophilized RNA extracted from *T. b. brucei* edited CPSF3^19^ was resuspended in nuclease-free water.

**Table 1:**
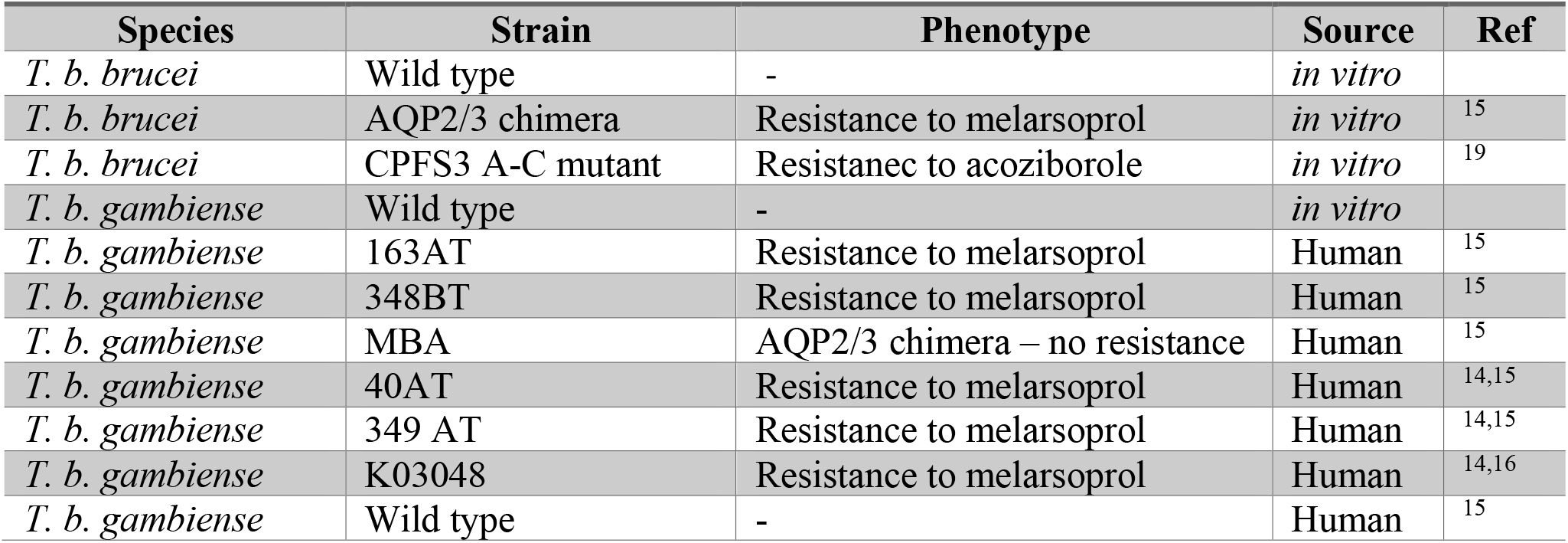
Trypanosome strains and isolates used in this study.

### *Lw*Cas13a enzyme expression and purification

The plasmid pC013-Twinstrep-SUMO-huLwCas13a (Addgene plasmid #90097) was used for the protein expression in *Escherichia coli* Rosetta^TM^ 2(DE3) pLysS competent cells ^25^. The protein expression and purification of *Leptotrichia wadei* Cas13a (*Lw*Cas13a) enzyme was performed as described in^27^ with slight modifications ^22^. TB medium was reconstituted by adding 47.8 g of TB powder to a 1-L flask, adding 8 mL of 100% (wt/vol) glycerol. Cell pellet was lysed with supplemented lysis buffer composed by 2 complete Ultra EDTA-free tablets (Roche), 500 mM NaCl, 100 mg lysozyme and 125-625 ng Deoxyribonuclease I from bovine pancreas (Sigma) to each 100 mL of lysis buffer. The *Lw*Cas13a was storage in single-use aliquots at -80°C to avoid freeze-thaw cycles.

### Design of RPA primers and crRNA

RPA primer pairs were designed using NCBI Primer-BLAST ^28^ using the custom parameters specified in^27^. For the wild-type CPSF3 SHERLOCK we use the reference sequence XM_839191.1 (Tb927.4.1340). For targeting the chimeric AQP2/AQP3_(814)_ the reference sequence KF564935.1 (*T. b. gambiense* strain 40AT) was used. The RPA primers used in the study are included in Supplementary Table 1. RPA forward primers include at the 5’ end, the T7 promoter sequence (5’-GAAATTAATACGACTCACTATAGGG-3’) that allow *in vitro* transcription of the amplified target by T7 polymerase.

The DNA templates used to generate the crRNAs consist of: 1) the target sequence (5’-3’) which will be the variable region between crRNA, also known as the spacer (with variable lengths from 20 to 28 nt), followed by 2) the direct repeat template common to all *Lw*Cas13a crRNAs (5’-GTTTTAGTCCCCTTCGTTTTTGGGGTAGTCTAGTCTAAATC-3’), followed by 3) the T7 promoter sequence at the 3’ end (5’-CCCTATATAGTGAGTCGTATTAATTTC-3’), which will allow *in vitro* transcription of the guides. DNA templates used are listed in Supplementary Table 1. All of the oligonucleotides mentioned were synthesized by ThermoFisher.

### *In vitro* transcription and purification of crRNAs

The crRNA synthesis was carried out using a DNA template and *in vitro* transcription mediated by T7 polymerase according to manufacturer’s instructions using the HiScribe™ T7 Quick High Yield RNA Synthesis Kit (NEB), as previously described ^27^. Briefly, we first annealed the crRNA DNA template and T7-3G oligonucleotide (5’- GAAATTAATACGACTCACTATAGGG-3’) by denaturation for 5 min followed by slow cooling ^22^. After *in vitro* transcription, the crRNAs were purified using magnetic beads (Agencourt RNAClean XP) and stored at 300 ng/μL in single-use aliquots at -80°C to avoid freeze-thaw cycles.

### SHERLOCK assay

The Specific High Sensitivity Enzymatic Reporter unLOCKing (SHERLOCK) assay was performed as described in^27^. The SHERLOCK reaction is a combination of a pre-amplification method by a reverse transcription (RT) recombinase polymerase amplification (RPA)^29^ and a specific RNA-target recognition by Cas13-CRISPR RNA-guide (crRNA) machinery (Abudayyeh et al., 2016). The RT-RPA was carried out using the TwistAmp Basic kit (TwistDx) following manufacturer’s instructions for 45 min at 42 °C as described by ^22^. The amplification step is followed by simultaneous *in vitro* transcription of the amplified target by T7 polymerase (Biosearch technology) and detection of the *Lw*Cas13a-crRNA using the same condition as described by ^22^. For this purpose, 4 μL of the RT-RPA product of each replicate was mixed with the following components in the following concentrations: 20 mM HEPES pH 6. 5, 9 mM MgCl2, 1 mM rNTP mix (NEB), 40 nM of *Lw*Cas13a, 2 U of Murine RNase inhibitor (NEB), 25 U of NxGen T7 RNA Polymerase (Biosearch technology), 25 nM of crRNA and 125 nM of RNaseAlert probe V2 (Invitrogen), in a final volume of 80 μL. The reaction was performed in 384-well plates, F-bottom, μClear bottom (Greiner) incubated at 37 °C in a TECAN plate reader (INFINITE F200 PRO M PLEX), in which 20 μL x 3 of each replicate reaction was distributed. Fluorescence measurements were collected every 10 minutes up to 3 hours. All SHERLOCK reactions were performed in triplicate and three fluorescence measurements were taken from each replicate. A negative control template (NCT) was added in parallel to each independent assay by supplementation nuclease-free water as input.

### Statistical analysis

The fluorescence intensity values obtained from the TECAN plate reader were analysed using Excel. For each triplicate reaction at each timepoint, the mean fluorescence intensity value was divided by the mean of the fluorescence intensity value of the NCT at the same timepoint, to obtain the fold-change over background fluorescence. The formula used is as follows:

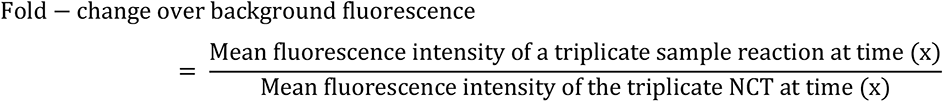

The graphs and statistical analysis were performed using GraphPad Prism (version 9.3.1) software. The Shapiro-Wilk normality test was performed to evaluate the type of data distribution. The non-parametric Mann-Whitney U test or the parametric bilateral unpaired t-test, with Welch’s correction, according to the data distribution results, were used for the statistical comparison with 95% confidence interval. For multiple comparisons, we applied the two-stage linear step-up procedure ^30^. Multiple sequence alignments were performed using Clustal Omega ^31^.

## Results

### Selection of SHERLOCK targets to detect the AQP2/AQP3_(814)_ chimera

We adapted our SHERLOCK4HAT ^22^ workflow for the detection of markers of resistance to melarsoprol through the formation of an *AQP2/3* chimera (Figure 1A). The *AQP2/3*_(814)_ chimera independently arose in two distinct HAT foci to currently made up 31.7 % of all known melarsoprol resistant HAT infections ^14,15^. The RPA forward primer amplified from nucleotide position 705 of the *AQP2* sequence, and the RPA reverse primer from position 841, which was specific to the region *AQP3* sequence of the chimera (Figure 1B). The sequences of the wild type (WT) *AQP2* and *AQP3* (Tb927.10.14170, Tb927.10.14160, respectively) and that of the *AQP2/3*_(814)_ chimera (KF564931.1) were aligned to select non-homologous regions between *AQP2* and *AQP3* genes for the new primers binding sites (Figure 1B). The crRNA guides were designed to target the region of the *AQP2/3*_(814)_ chimera that was analogous to *AQP2* (Figure 1B). We evaluated three crRNA for the ability to discriminate between RNAs extracted from *in vitro* derived cells where both *AQP2* and *AQP3* were knocked out and the *AQP2/3_(814_*_)_ chimera was expressed from the rRNA intergenic region ^32^, and from wild type *T. b. gambiense* or *T. b. brucei* cells (Figure 1C). The three crRNAs specifically detected the cells expressing the *AQP2/3_(814)_* chimera and did not cross-react with the WT trypanosomes or human cells (Figure 1C).

**Figure 1.**
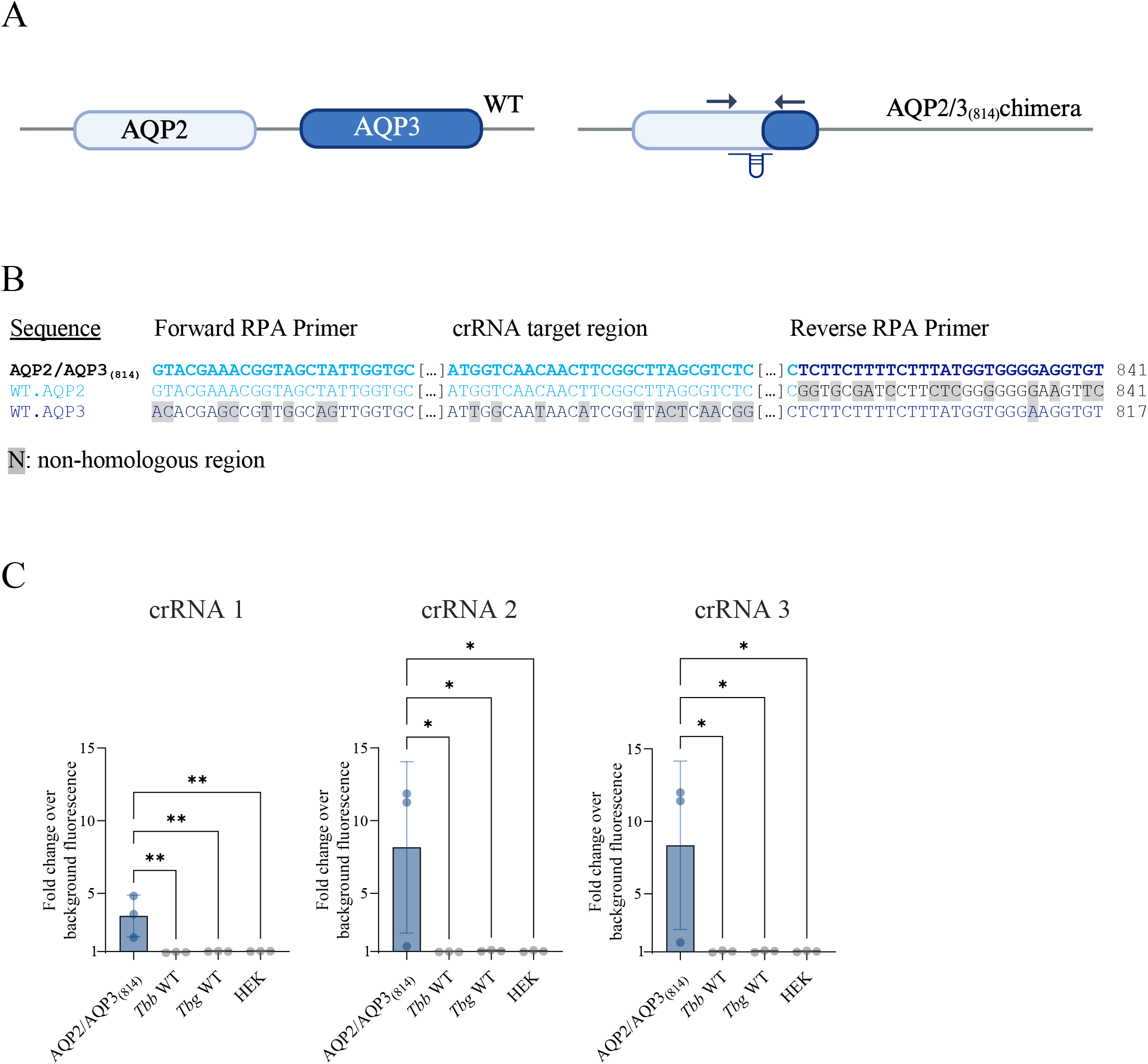
Detection of the *AQP2/3_(814)_* chimera by SHERLOCK. (A) Schematic of the *AQP2* and *AQP3* wild type locus (left) and the *AQP2/AQP3_(814)_* chimera (right). The RPA primers (arrows) and crRNA target (RNA guide) regions for detection of the *AQP2/AQP3_(814)_* chimera are shown. (B) Alignments of the chimera *AQP2/AQP3_(814)_* (KF564931.1), WT *AQP2* (Tb927.10.14170) and *AQP3* (Tb927.10.14160) sequences are shown with the non- homologous regions highlighted in gray. (C) crRNA screening for the detection of the *AQP2/AQP3_(814)_* chimera using RNA from in vitro derived *AQP2/AQP3_(814)_*, WT *T.b.b.* (*T. b. brucei*), WT *T.b.g.* (*T. b. gambiense*), and human embryonic kidney (HEK) cells. RNA input at 1 ng/μL. Asterisk represents p values at (*) p<0.001 and (**) p<0.0001.

### The AQP2/AQP3_(814)_-specific SHERLOCK assay can be used to identify samples from cases of HAT relapses

After demonstrating that the AQP2/AQP3_(814)_-specific SHERLOCK assay was able to detect RNA from *in vitro* modified cells, we wanted to investigate whether it could also detect AQP2/AQP3_(814)_-chimeric RNAs of parasites isolated from patients, from two geographically distinct locations, showing relapses following treatment with melarsoprol in the Mbuji-Mayi region of the Democratic Republic of the Congo and Mundri County in South Sudan (Figure 2A), which have been associated with mutations in the *AQP2/3* genes^14–16^ (Figure 2B). We tested 4 isolates: (1) a WT strain from a patient infected with *T. b. gambiense* with no mutation in the *AQP2/3* locus and no relapse; (2) the 163AT, 40AT, 349AT and K03048 strains from patients with HAT relapse infected with *T. b. gambiense* bearing the AQP2/AQP3_(814)_ mutation^14–16^; (3) the MBA strain from a patient with unknown treatment outcome, infected with *T. b. gambiense* containing an *AQP2*/3 chimera with the first 677 bp from *AQP2*, 202 bp of *AQP3* and the last 60 bp from *AQP2*, termed as *AQP2/3*_(678–880)_; and the 348BT strain isolated from a cured patient that contains both *AQP2/3*_(814)_ and *AQP2/3*_(880)_ (first 879 bp from *AQP2*, a point mutation at T869C and the last 60 bp from *AQP3*) chimeras (Figure 2B)_15._

**Figure 2.**
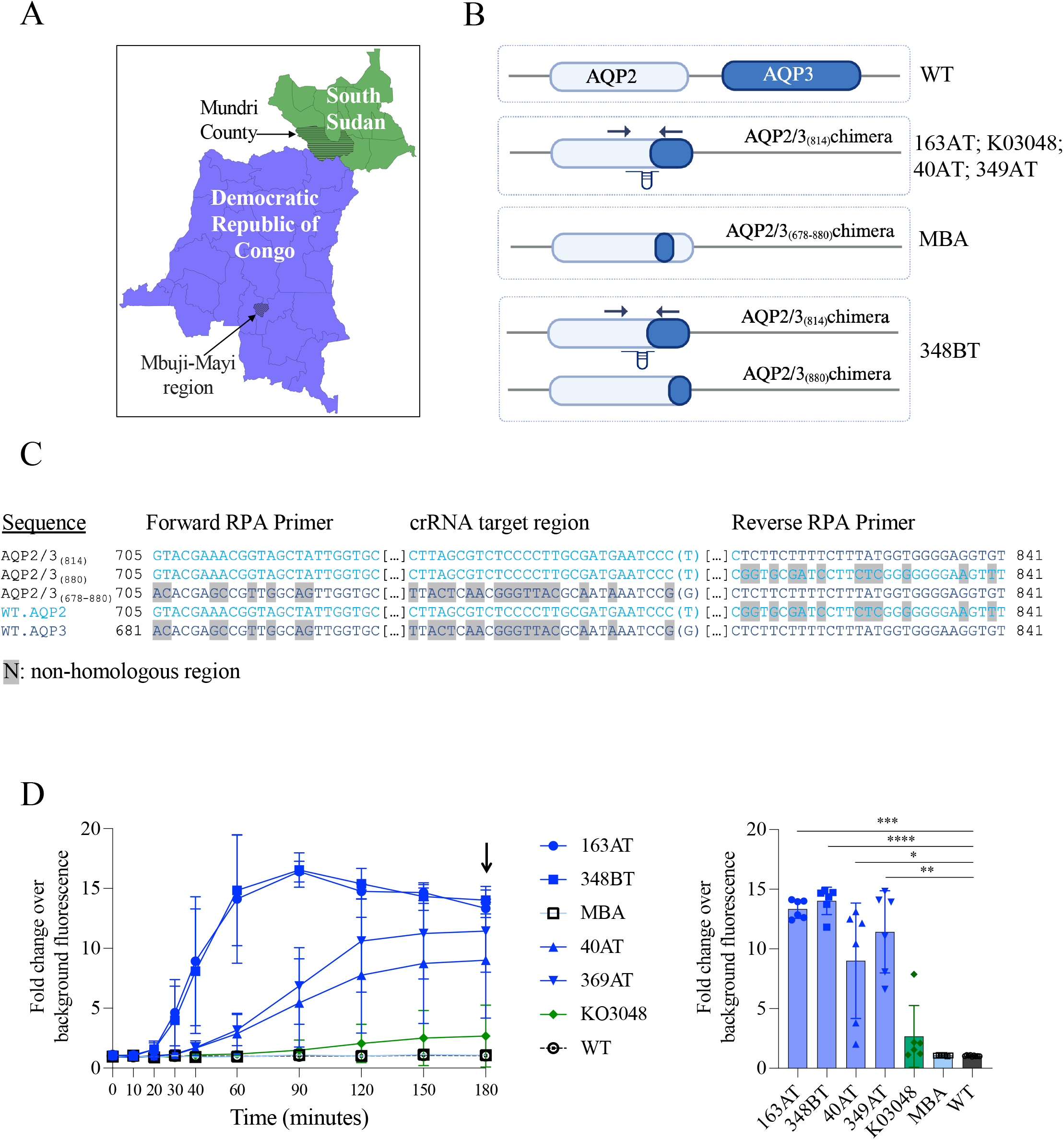
Evaluation of the *AQP2/AQP3_(814)_* SHERLOCK assay on isolates from HAT patients with relapses associated with the formation of *AQP2/3* chimeras. (A) Map showing the regions where patient with relapse after treatment was detected (B) Schematics of the *AQP2* and *AQP3* loci in 4 different strains isolated from patients in the DRC (WT, 163AT, MBA and 348BT) and South Sudan (K03048). The RPA primers (arrows) and crRNA target (RNA guide) regions for detection of the *AQP2/AQP3_(814)_* chimera are shown. (C) Alignments of the WT *AQP2* (Tb927.10.14170) and *AQP3* (Tb927.10.14160); *AQP2/AQP3_(814)_* (KF564931.1); *AQP2/3*_(880)_ (KM282050) and *AQP2/3*_(678-880)_ (KM282034) sequences are shown with the non- homologous regions highlighted in gray. (D) Kinetic representation of the Cas13a reaction using the A/3_(814)_-specific SHERLOCK assay testing the different RNAs extracted from the 6 strains isolated from DRC patients (163AT, 348BT, 40AT, 349AT, MBA and WT) and one strain form South Sudan (K03048) (left graph). Representation of the fold-change over background fluorescence after 180 min (time point taken indicated by black arrow in kinetic plot) of Cas13a reaction using the A/3_(814)_-specific SHERLOCK assay analysing the RNAs from the different isolated strains (right graph).

The sequences of the wild type AQP2 and 3, the *AQP2/3*_(814)_ chimera (KF564931.1), *AQP2/3*_(880)_ (KM282050) and *AQP2/3*_(678-880)_ (KM282034) were aligned to select non-homologous regions between *AQP2* and *AQP3* genes to design a second RPA primer pair (Figure 2B, Supplementary table 1): the selected RPA forward primer amplified from nucleotide position 730 of the *AQP2* sequence, and the RPA reverse primer from position 829 of the chimera sequence corresponding to the *AQP3* sequence. Our crRNA guides should not target the *AQP2/3*_(880)_ (KM282050) or *AQP2/3*_(678-880)_ (KM282034) chimeras (Figure 2B and C). We first screened the samples using two pairs of RPA primer and two crRNA guides (Supplementary figure 1A). The crRNA2 and RPA primer pair 1 were selected to assess the samples and this SHERLOCK assay was shown to accurately discriminate between patient samples that contained the *AQP2/3*_(814)_ chimera or not (Figure 2D). As expected, the 163AT, 40AT, 349AT and 348 BT samples showed a robust signal in the SHERLOCK *AQP2/3*_(814)_ assay. The K03048 sample from South Sudan scored as positive in the assay, yet it showed a lower fold-change, likely because the amount of RNA extracted from this sample was low (Figure 2D). These results confirm that the SHERLOCK *AQP2/3*_(814)_ assay can detect the specific mutation associated with melarsoprol resistance from patient samples.

### SHERLOCK detection of a single nucleotide variant in the *CPSF3* gene

Taking advantage of the specificity of SHERLOCK, we developed an assay that can discriminate between the WT and the *in vitro* derived single nucleotide variant (SNV) of the *CPSF3* gene (*CPSF3_(SNV)_,* accession number XM_839191: N^232^H, AAT-CAT), hereon referred to as *CPSF3_(SNV)_* which confers resistance to acoziborole ^19,33^. We first screened RPA primer pairs that were either 30 nt or 25 nt long (Supplementary figure 1B) and designed to amplify the region containing the SNV. The RPA primer pair target amplification was assessed using four different crRNA guides specific for the detection of *CPSF3_(SNV)_*. We observed higher fold-change values with the 30 nt RPA primer pair as compared to the 25 nt primer pair (Supplementary Figure 1B). Therefore, we selected the long RPA primer pair for subsequent optimization of the SHERLOCK assay.

Then, 12 crRNA guides were screened for detection of the *CPSF3_(SNV)_*. We designed crRNAs using the following criteria: 1) setting the SNV complementary base at position 3 (counting from the 3’ end of the crRNA) and the artificial mismatch (if any) at position 5; and 2) setting the SNV complementary base at position 6 and the mismatch (if any) at position 4 ^23,25,34^ (Figure 3A). In addition, we designed crRNAs with spacer lengths shorter than described as optimal for *Lw*Cas13a (28 nt, ^27^). Indeed, reducing the spacer length to a maximum of 20 nt reduced the enzyme activity but maintained, or even improved, its ability to discriminate single mismatches by reducing background fluorescence in the off-target sample ^25,34,35^ (Figure 3B). Finally, since consecutive double substitutions in the spacer have been described as effective in the loss of collateral cleavage activity of the Cas13a enzyme ^35^, we also evaluated the efficacy of placing the synthetic mismatch adjacent to our SNV, in order to generate a double mismatch in the off-target sequence (Figure 3B).

**Figure 3.**
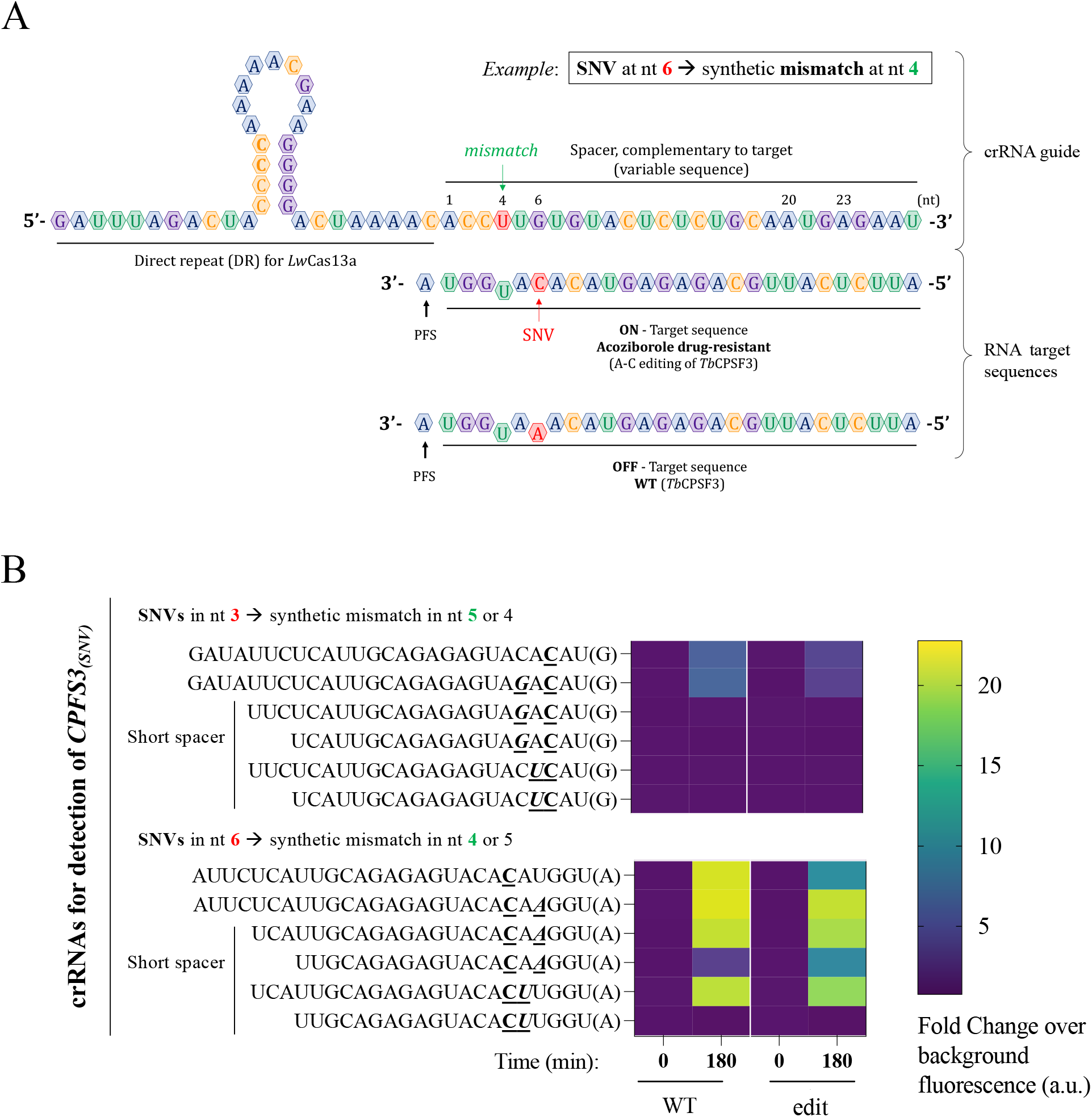
**Optimization of a SHERLOCK assay for the detection of *CPSF3* single nucleotide variant (SNV)**. (A) Schematic of the crRNAs design for the detection of the single nucleotide variant (SNV) in the *CPSF3* gene. Position of the SNV in red and the inclusion of the synthetic mismatch along the crRNA spacer in green. (B) Screening of crRNAs designed for the detection of *CPSF3*-edited cells. The target sequence of each crRNA is indicated, as well as the PFS in brackets. crRNAs used for detection of the *CPSF3_(SNV)_*: 1) SNV fixed at position 3, and 2) SNV fixed at position 6. SNV is in underlined and bold character, and synthetic mismatch is in underlined, bold and italicized character. crRNAs with different spacer lengths were evaluated (20, 23 and 27 nt). The heat map shows the fold change over background fluorescence intensity values collected at time zero and after 3 h of *Lw*Cas13a detection reaction. G, guanine; A, adenine; U, uridine; C, cytosine; PFS, protospacer flanking site; *Tb*CPSF3, *Trypanosoma brucei gambiense* cleavage and polyadenylation factor 3; WT, wild type.

The crRNAs designed to detect the SNV at position 3 showed lower fluorescence emission and were undetectable when using the crRNAs with a spacer length of 20 or 23 nt (Figure 3B). In contrast, the crRNAs detecting the SNV at position 6 gave high fluorescence intensity values with both WT and *CPSF3_(SNV)_*. The fluorescence was reduced only when the length of the spacer was limited to 20 nt (Figure 3B). The best performing crRNA guide contained the SNV complementary base at position 6 and a synthetic mismatch at position 4, with a spacer length of 20 nt (Figure 3B). We then evaluated whether the nucleotide selected as the artificial mismatch could improve the specificity of the SHERLOCK *CPSF3_(SNV)_* assay. We selected the best candidate obtained in the first screening (Figure 3B), which used an uracil (U) as the mismatch (Figure 4A), as well as two other crRNAs, one using cytosine (Figure 4B) and the other guanine (Figure 4C) as the artificial mismatch. Using either an uracil or a guanine base as mismatch, the SHERLOCK assays showed statistically significant difference in fluorescence intensity when comparing total RNAs from *CPSF3_(SNV)_* to WT RNA (Figure 4A and C). Using a guanine, the fluorescence from the off-target (WT) sequence was reduced to near background (Figure 4C). These data show that the SHERLOCK assay can be used to detect SNV in *T. brucei* cells resistant to acoziborole.

**Figure 4.**
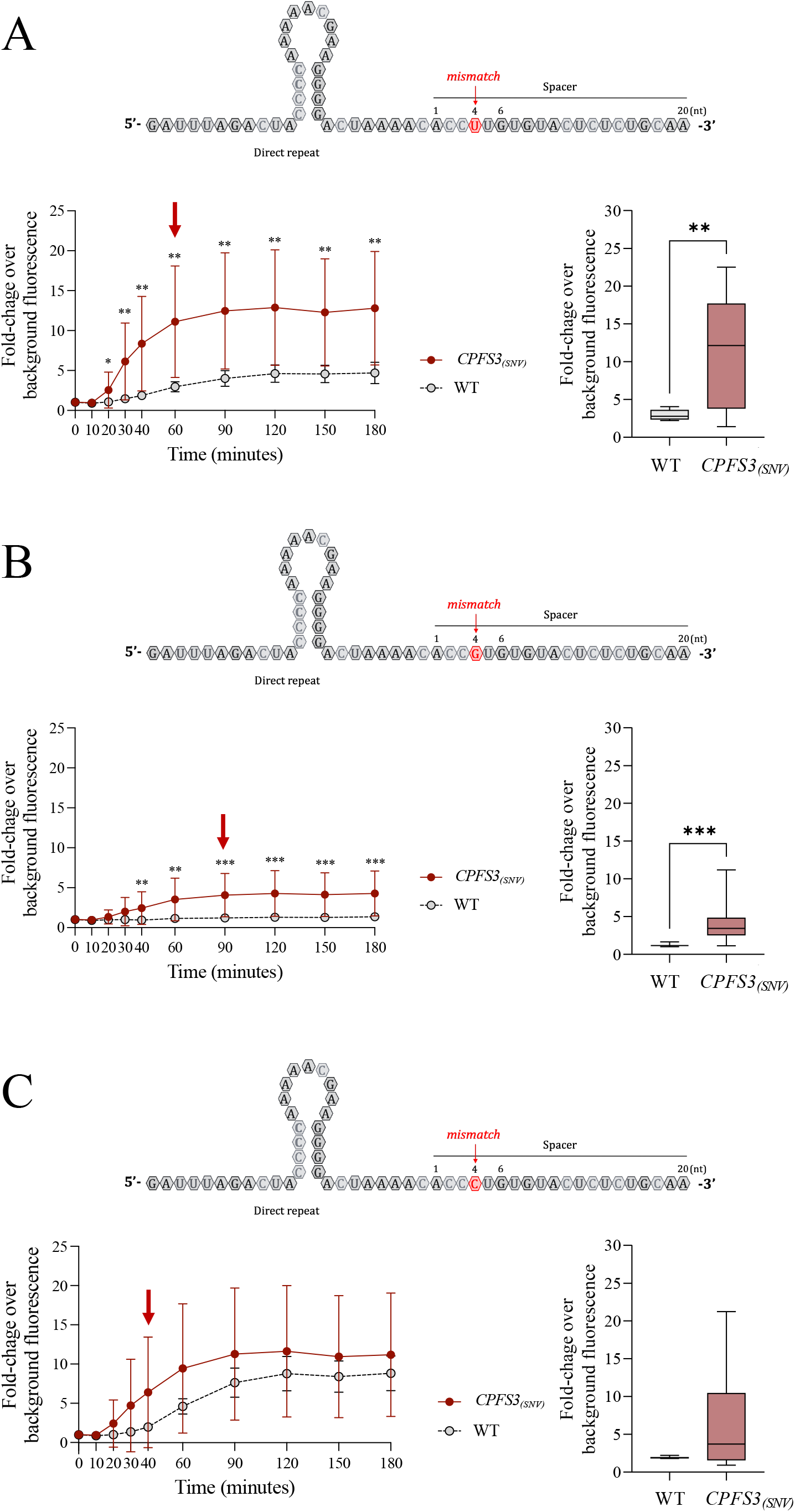
Evaluation of selected nucleotides used as synthetic mismatches in the *CPSF3_(SNV)_* SHERLOCK assay. Kinetic representation of *Lw*Cas13a reaction for the selected base for the synthetic mismatch at nt 4: uracil (A), guanine (B) or cytosine (C). The fold-change over background fluorescence was indicated at different time points of the *Lw*Cas13a reaction. Results from WT cells are shown with open circles and those from edited *CPSF3_(SNV)_* cells with red dots. Box-plots represent fold-change over background fluorescence intensity values using RNA from WT cells (gray box-plots) or RNA from *CPSF3_(SNV)_*-edited cells (red box-plots) at the selected time-point, here indicated by a red arrow in each kinetic plot. RNA input at 5 ng/μL. The p values are represented by asterisks as follows: p<0.05 (*), p<0.01 (**) and p<0.001 (***).

## Discussion

Here we describe the development of new drug resistance SHERLOCK assays for HAT that could be used for epidemiological surveillance. There are currently no molecular diagnostics that can screen for emerging drug resistance in HAT patients, yet resistance to all the currently approved drugs used to treat HAT has been detected either in the field or can be generated in a laboratory. The two SHERLOCK assays described here can distinguish WT and either gene chimera’s or SNV targets. As an RNA diagnostic, these SHERLOCK assays can be used to screen patients presenting relapse after treatment and could be used to discriminate between relapse or reinfection, which is critical for adapting drug treatment.

Although no longer used for the treatment of gHAT, melarsoprol is still recommended by the WHO for treatment of rHAT at stage 2 in children or as a second option in adults. Unlike gHAT, rHAT is an acute infection, with disease progression taking from weeks to months. Resistance to melarsoprol would, therefore, represent a significant obstacle to control rHAT. In this study, we have developed a SHERLOCK assay targeting mutations in the *AQP2/3* locus that were initially identified in circulating parasites by sequencing samples from patients with relapses after treatment^14,15^ . Our high-throughput AQP2/3_(814)_ SHERLOCK assay detected all parasites involved in relapses without cross reactivity and could be quickly implemented in reference labs in the field. One limitation of using SHERLOCK for detecting drug resistance is the intrinsic dependency of the method on prior knowledge of the gene mutations for the design of RPA primers and crRNAs. However, by using our existing SHERLOCK4HAT workflow ^22^, the assay could rapidly be adapted and implemented once a mutation has been identified.

Acoziborole will likely become the next frontline drug for the treatment of gHAT to move towards elimination of the disease by 2030 ^17,36^. Like for the other drugs used to treat HAT, resistance to acoziborole can be generated in the laboratory through a single point mutation in the *CPSF3* gene ^19,33^. Several SHERLOCK assays have been developed to detect SNV ^23,25,34^, and *Lw*Cas13a, used in this study, is capable of tolerating mismatches, yet with a reduced cutting efficient ^37^. This could be ameliorated by optimising the crRNA guide length, the positioning of the mismatch, and/or the Cas13 variant used ^34^. In our *CPSF3_(SNV)_* SHERLOCK assay, we found that the most efficient combination was a 20 nt crRNA, the SNV at position 6 and the synthetic mismatch at position 4. Within the *CPSF3_(SNV)_* SHERLOCK assay, there is still scope for optimisation to improve the specificity of the assay and which Cas13 variant could provide better on vs. off target discrimination. In our assays, the nucleotide selected for the mismatch had the most profound effect on the sensitivity of the assay, especially in the discrimination between on and off target. Here, the use of a guanine as the mismatch at position 4 showed the best discrimination between the two targets. Should resistance arise from a SNV or a chimeric gene formation, fully exploiting how the Cas13a-RNA complex is formed will be key to developing a robust assay for epidemiological surveillance.

This study demonstrates the versatility of the SHERLOCK technology to detect known genetic modifications directly associated to drug resistance, using existing workflows. The major challenge to develop a SHERLOCK assay is that the targeted genetic mutation must be both known and stable. However, for rHAT treatment with melarsoprol, we showed that known resistance mechanisms through the formation of chimeras, that have arisen in geographically separated regions, are suitable targets for SHERLOCK. Given the adaptability of SHERLOCK, generating a HAT antimicrobial resistance toolbox will be an invaluable asset for epidemiological surveillance to screen any cases of relapse. This will be critical not only to track the incidence of resistance, but also for subsequent selection of the most individually adapted drug treatment.

## Data Availability

All data produced in the present study are available upon reasonable request to the authors

## Abbreviations

AQP: Aquaglyceroporin
CPSF3: Cleavage and polyadenylation specific factor 3
CRISPR: Clustered regularly interspaced short palindromic repeats crRNA: CRISPR RNA
HAT: Human African trypanosomiasis HEK: Human embryonic kidney
*Lw*Cas13a: *Leptotrichia wade*i CRISPR-associated protein 13 a NCT: Negative control template
NECT: Nifurtimox- eflornithine combination therapy PAM: Protospacer adjacent motif
RPA: Recombinase polymerase amplification RT: Reverse transcription
SHERLOCK: Specific High-sensitivity Enzymatic Reporter unlocking SNV: Single nucleotide variant
SUMO: Small ubiquitin-like modifier WT: Wild-type

## Acknowledgments

The authors acknowledge the support of the researchers who made this work possible by providing the necessary genetic materials: *T. b. gambiense* ELIANE cell pellets were provided by Pr. Annette MacLeod (University of Glasgow, UK); lyophilized RNA extracted from *CPSF3*-edited *T. b. brucei* were provided by Pr. David Horn (University of Dundee, UK); RNA from chimeric AQP2/AQP3 *T. b. gambiense* isolates were provided by Dr. Nick Van Reet (Institute of Tropical Medicine, Antwerp) and Pr. Pascal Maeser (Swiss Tropical and Public Health Institute, Switzerland).

## Author contribution

EPA, BR and LG conceived and designed the experiments. EPA and ADH performed the experiments. EPA, BR and LG analysed the data. BR and LG contributed reagents, materials and analysis tools. EPA, BR and LG wrote the paper.

---

**Supplementary** Figure 1**. RPA primer screening.** (A) Representation of the results obtained in the screening of RPA primers (1 and 2) and crRNAs (2 and 3) for the AQP2/3_(814)_-specific SHERLOCK assay using as input the different RNAs from the extraction of strains isolated from gHAT patients that contains the AQP2/3_(814)_ (163AT and 384BT) or not (MBA and WT). (B) Evaluation of the pairs of RPA longer length primers (30 nt) and shorter length primers (25 nt) by SHERLOCK assay using four potential crRNAs (1 to 4) specific for the detection of genetic material of *CPSF3_(SNV)_-*edited cells. The data were represented as the fold-change over background fluorescence, taking the values obtained after 60 min of *Lw*Cas13a detection reaction.

**Supplementary Table 1**. RPA primers and crRNA templates used in this study.

## References

1. WHO. Global report on neglected tropical diseases 2024. 2024.

2. WHO. Guidelines for the treatment of human African trypanosomiasis. *WHO* 2024.

3. Dickie EA, Giordani F, Gould MK, et al. New Drugs for Human African Trypanosomiasis: A Twenty First Century Success Story. Trop Med Infect Dis 2020; 5(1).

4. Checchi F, Chappuis F, Karunakara U, Priotto G, Chandramohan D. Accuracy of five algorithms to diagnose gambiense human African trypanosomiasis. PLoS Negl Trop Dis 2011; 5(7): e1233.

5. Lindner AK, Lejon V, Chappuis F, et al. New WHO guidelines for treatment of gambiense human African trypanosomiasis including fexinidazole: substantial changes for clinical practice. Lancet Infect Dis 2020; 20(2): e38–e46.

6. Deeks ED. Fexinidazole: First Global Approval. Drugs 2019; 79(2): 215–20.

7. Pelfrene E, Harvey Allchurch M, Ntamabyaliro N, et al. The European Medicines Agency’s scientific opinion on oral fexinidazole for human African trypanosomiasis. PLoS Negl Trop Dis 2019; 13(6): e0007381.

8. Wyllie S, Foth BJ, Kelner A, Sokolova AY, Berriman M, Fairlamb AH. Nitroheterocyclic drug resistance mechanisms in Trypanosoma brucei. J Antimicrob Chemother 2016; 71(3): 625–34.

9. Barrett MP, Vincent IM, Burchmore RJ, Kazibwe AJ, Matovu E. Drug resistance in human African trypanosomiasis. Future Microbiol 2011; 6(9): 1037–47.

10. Baker N, de Koning HP, Maser P, Horn D. Drug resistance in African trypanosomiasis: the melarsoprol and pentamidine story. Trends Parasitol 2013; 29(3): 110–8.

11. Fairlamb AH, Horn D. Melarsoprol Resistance in African Trypanosomiasis. Trends Parasitol 2018; 34(6): 481–92.

12. Baker N, Glover L, Munday JC, et al. Aquaglyceroporin 2 controls susceptibility to melarsoprol and pentamidine in African trypanosomes. Proc Natl Acad Sci U S A 2012; 109(27): 10996–1001.

13. Bassarak B, Uzcategui NL, Schonfeld C, Duszenko M. Functional characterization of three aquaglyceroporins from Trypanosoma brucei in osmoregulation and glycerol transport. Cell Physiol Biochem 2011; 27(3-4): 411–20.

14. Graf FE, Ludin P, Wenzler T, et al. Aquaporin 2 mutations in Trypanosoma brucei gambiense field isolates correlate with decreased susceptibility to pentamidine and melarsoprol. PLoS Negl Trop Dis 2013; 7(10): e2475.

15. Pyana Pati P, Van Reet N, Mumba Ngoyi D, Ngay Lukusa I, Karhemere Bin Shamamba S, Buscher P. Melarsoprol sensitivity profile of Trypanosoma brucei gambiense isolates from cured and relapsed sleeping sickness patients from the Democratic Republic of the Congo. PLoS Negl Trop Dis 2014; 8(10): e3212.

16. Maina NW, Oberle M, Otieno C, et al. Isolation and propagation of Trypanosoma brucei gambiense from sleeping sickness patients in south Sudan. Trans R Soc Trop Med Hyg 2007; 101(6): 540–6.

17. Betu Kumeso VK, Kalonji WM, Rembry S, et al. Efficacy and safety of acoziborole in patients with human African trypanosomiasis caused by Trypanosoma brucei gambiense: a multicentre, open-label, single-arm, phase 2/3 trial. Lancet Infect Dis 2023; 23(4): 463–70.

18. Tarral A, Hovsepian L, Duvauchelle T, et al. Determination of the Optimal Single Dose Treatment for Acoziborole, a Novel Drug for the Treatment of Human African Trypanosomiasis: First-in-Human Study. Clin Pharmacokinet 2023; 62(3): 481–91.

19. Wall RJ, Rico E, Lukac I, et al. Clinical and veterinary trypanocidal benzoxaboroles target CPSF3. Proc Natl Acad Sci U S A 2018; 115(38): 9616–21.

20. Sharma A, Cipriano M, Ferrins L, Hajduk SL, Mensa-Wilmot K. Hypothesis-generating proteome perturbation to identify NEU-4438 and acoziborole modes of action in the African Trypanosome. iScience 2022; 25(11): 105302.

21. Kovarova J, Novotna M, Faria J, et al. CRISPR/Cas9-based precision tagging of essential genes in bloodstream form African trypanosomes. Mol Biochem Parasitol 2022; 249: 111476.

22. Sima ND-H, A.; Perlaza, BL.; Ungeheuer, MN.; Rotureau, B.; Glover, L. SHERLOCK4HAT: a CRISPR-based tool kit for diagnosis of Human African Trypanosomiasis. medRxiv 2022.

23. Cunningham CH, Hennelly CM, Lin JT, et al. A novel CRISPR-based malaria diagnostic capable of Plasmodium detection, species differentiation, and drug-resistance genotyping. EBioMedicine 2021; 68: 103415.

24. Gootenberg JS, Abudayyeh OO, Kellner MJ, Joung J, Collins JJ, Zhang F. Multiplexed and portable nucleic acid detection platform with Cas13, Cas12a, and Csm6. Science 2018; 360(6387): 439–44.

25. Gootenberg JS, Abudayyeh OO, Lee JW, et al. Nucleic acid detection with CRISPR- Cas13a/C2c2. Science 2017; 356(6336): 438-42.

26. Myhrvold C, Freije CA, Gootenberg JS, et al. Field-deployable viral diagnostics using CRISPR-Cas13. Science 2018; 360(6387): 444-8.

27. Kellner MJ, Koob JG, Gootenberg JS, Abudayyeh OO, Zhang F. SHERLOCK: nucleic acid detection with CRISPR nucleases. Nat Protoc 2019; 14(10): 2986–3012.

28. Ye J, Coulouris G, Zaretskaya I, Cutcutache I, Rozen S, Madden TL. Primer-BLAST: a tool to design target-specific primers for polymerase chain reaction. BMC Bioinformatics 2012; 13: 134.

29. Piepenburg O, Williams CH, Stemple DL, Armes NA. DNA detection using recombination proteins. PLoS Biol 2006; 4(7): e204.

30. Benjamini YaH, Y. Controlling the False Discovery Rate: a Practical and Powerful Approach to Multiple Testing. *The Journal of the Royal Statistical Society*, Series B (Statistical Methodology) 1995; 57(1): 289–300.

31. Sievers F, Higgins DG. Clustal Omega for making accurate alignments of many protein sequences. Protein Sci 2018; 27(1): 135–45.

32. Graf FE, Baker N, Munday JC, de Koning HP, Horn D, Maser P. Chimerization at the AQP2-AQP3 locus is the genetic basis of melarsoprol-pentamidine cross-resistance in clinical Trypanosoma brucei gambiense isolates. Int J Parasitol Drugs Drug Resist 2015; 5(2): 65–8.

33. Altmann S, Rico E, Carvalho S, et al. Oligo targeting for profiling drug resistance mutations in the parasitic trypanosomatids. Nucleic Acids Res 2022; 50(14): e79.

34. Molina Vargas AM, Sinha S, Osborn R, et al. New design strategies for ultra-specific CRISPR-Cas13a-based RNA detection with single-nucleotide mismatch sensitivity. Nucleic Acids Res 2024; 52(2): 921–39.

35. Abudayyeh OO, Gootenberg JS, Essletzbichler P, et al. RNA targeting with CRISPR- Cas13. Nature 2017; 550(7675): 4280-4.

36. Franco JR, Priotto G, Paone M, et al. The elimination of human African trypanosomiasis: Monitoring progress towards the 2021-2030 WHO road map targets. PLoS Negl Trop Dis 2024; 18(4): e0012111.

37. Mantena S, Pillai PP, Petros BA, et al. Model-directed generation of CRISPR-Cas13a guide RNAs designs artificial sequences that improve nucleic acid detection. bioRxiv 2023.

