## Supplementary figures and images for "A next generation CRISPR diagnostic tool to survey drug resistance in Human African Trypanosomiasis"

### Supplementary figure 1

Supplementary Figure 1

A

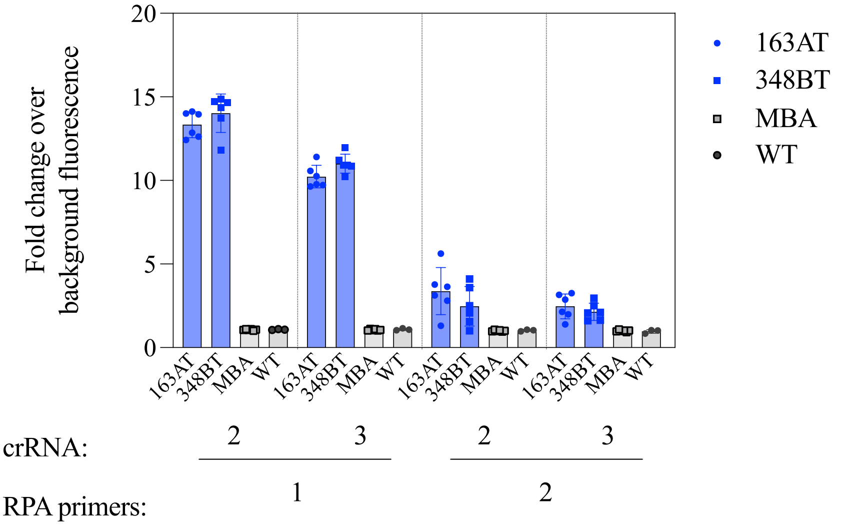


B

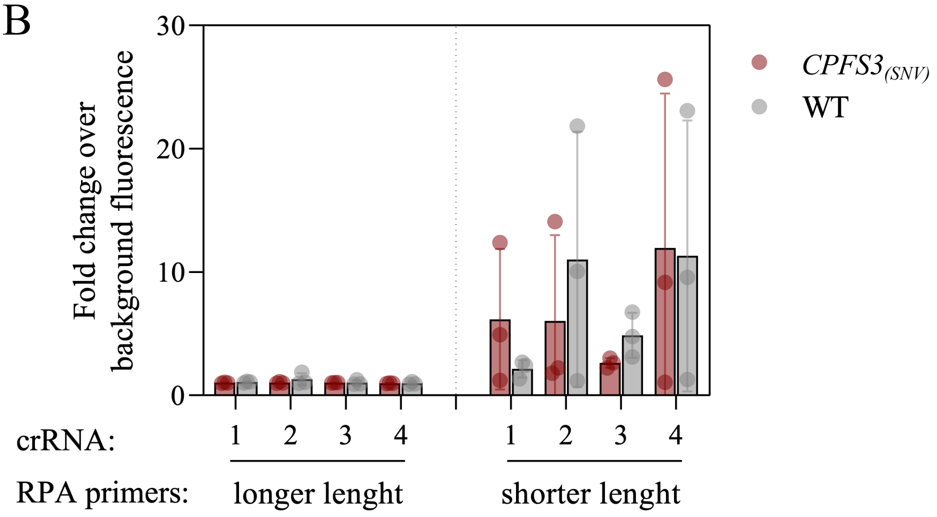
